# Assessing quadriceps strength in patellofemoral pain patients: A study on the reliability and validity of a low-cost load-cell for clinical practice

**DOI:** 10.1101/2024.02.01.24301977

**Authors:** Germari Deysel, Mariaan van Aswegen, Mark Kramer

## Abstract

**Background:** Patellofemoral pain (PFP) is a common knee complaint affecting diverse populations both acutely and chronically. Quadriceps muscle weakness is one possible aetiology, but current devices for measuring muscle strength (isokinetic dynamometer and hand-held dynamometers) are frequently too expensive for practitioners, especially in under-resourced settings. There is a need to evaluate a low-cost device to manage rehabilitation of people with PFP.

**Methods:** Isometric quadriceps strength of participants aged 18-35 years (total [n = 33], control group [n = 17] and PFP group [n = 16]) were evaluated on an isokinetic dynamometer and a low-cost load cell at baseline and after an 8-week non-standardised intervention for validity scores.

**Results:** The load cell showed high absolute and relative reliability (intraclass correlation coefficient = 0.89-0.99; typical error of measurement = 3.9-10.4%). Clinically meaningful difference scores (12.2-45 Nm) were greater than the typical error of measurement, implying sufficient sensitivity of the load cell to measure true changes in isometric quadricep strength. Strong to very strong correlations were evident between the load cell and isokinetic dynamometer torque measurements (r = 0.88-0.90, SEE = 0.05-0.07 Nm), but slope values (β = 0.65-0.77) indicated that torque from the load cell was typically lower than that obtained from the isokinetic dynamometer. An average systematic bias of 16.3-28.8 Nm was evident in favour of the isokinetic dynamometer, with no statistically significant between-group differences being noted between the baseline and follow-up testing.

**Conclusion:** The load cell is a reliable instrument, sensitive enough to detect clinically meaningful differences in quadriceps strength in healthy individuals and those with PFP. The load cell lacks validity and cannot replace isokinetic dynamometry. Given the low cost and excellent reliability, the load cell can be a valuable tool to assess quadriceps muscle deficits and track rehabilitation progress in people with PFP.

## Introduction

Patellofemoral pain (PFP) is a common knee complaint characterised by retropatellar pain (pain behind the kneecap) or peripatellar pain (pain around the kneecap), aggravated by at least one knee loading activity during weight bearing on a flexed knee (e.g., squatting, stair ambulation, jogging, running, hopping or jumping) [1]. Prevalence rates of PFP vary according to age, gender, and activity levels with the literature showing rates of 28% in adolescents [2], 23% in adults [3], 15% in female adults [4] and 40% in recreational runners [5]. People living with PFP often struggle with acute and chronic effects such as physical, emotional and social problems during sports participation, activities related to work and activities of daily living, which can persist several years [6–10]. Furthermore, PFP is considered a risk factor for the development of patellofemoral osteoarthritis [11], which has been attributed to reduced quadriceps strength relative to task-related loading of the patellofemoral joint [5,12,13]. It is therefore plausible to consider quadriceps strength as a *protective agent* against patellofemoral osteoarthritis cartilage loss [14].

Given that people with PFP can be divided into different subgroups, depending on the individual aetiologies [15], it is important to determine whether quadriceps weakness is part of the cause for PFP in the individual, and to track whether progress is made when conducting an exercise program to improve quadriceps strength [15]. Presently, the ‘gold standard’ instrument for the evaluation of quadriceps strength is the isokinetic dynamometer (ID) [16,17], which is beyond the affordability of most clinicians (∼R500k-R1.50mil). Although a more reasonable alternative to the ID exists, such as a handheld dynamometer (HHD) [17], the reliability of an HHD depends on the strength and aptitude of the practitioner [18,19]. The latter is especially true when assessing stronger individuals or muscle strength around larger joints such as the knee joint [17] which may have important clinical implications for both patients and practitioners. Furthermore, HHDs are still relatively expensive (∼R20k) when considering starting a rehabilitation practice, especially in under-resourced communities and clinical settings. Therefore, the purpose of this study was to: (i) establish the validity and reliability of a low-cost load cell (LC) (∼R2k) for the assessment of isometric quadriceps muscle strength in people with PFP, (ii) evaluate the clinically meaningful difference (CMD) needed to enhance practitioner-based decision-making, and (iii) assess differences in isometric quadriceps strength with the use of a low-cost device between those with PFP and healthy control following an 8-week intervention.

## Materials and Methods

### Study design

This study used a repeated measures mixed study design consisting of a combination of between and within subject factors [20]. The between-subjects independent variable was group allocation (i.e., control group and PFP group), where group assignment was not randomised (due to presence/absence of PFP). Within-subjects independent variables included time (repeated measures) and extremity (injured [or non-dominant] and uninjured [or dominant] limb).

### Participants

A total of 35 participants volunteered for the study, of which 17 were part of the control group (female [n=10] and male [n=7]) and 18 were part of the PFP group (female [n=15] and male [n=3]. Given the requirements of the study design, a minimum total sample size of 24 was calculated based on a repeated measures analysis of variance (ANOVA) design that incorporated (i) a within-between interaction, (ii) a moderate effect size (f = 0.25), (iii) a type-I error rate of 5% (α = 0.05), (iv) a type-II error rate of 20% (β = 0.20), (v) 2 groups (control [n = 17] and PFP [n = 17]), (vi) 2 repeated measurements, (vii) an anticipated dropout of 10%, and (viii) a minimum expected correlation of 0.50 among repeated measurements [21]. Accounting for a potential drop-out of 20%, the minimum sample size for adequate statistical power was 29 participants. Two participants were lost to follow-up, both in the PFP group (female [n=1] and male [n=1]), resulting in a final sample size of 33 participants.

Participants were recruited by dispersing electronic flyers via social media platforms (e.g., Facebook, WhatsApp), inviting prospective participants to contact the researcher for an information letter which outlined the details of the study. Participants who were willing to take part in the study were screened for eligibility (see inclusion and exclusion criteria below) and received an informed consent form to read through and were given 72 hours to sign and submit the form to an independent person affiliated with the study.

The inclusion criteria for the PFP group of this consisted of the following: (i) aged between 18 and 35 years, (ii) could be male or female, (iii) had to have retropatellar and/or peripatellar pain aggravated by at least one activity that loads the patellofemoral joint during weight bearing on a flexed knee, such as squatting, stair climbing, jogging/running, and hopping/jumping, and (iv) had to participate in some form of rehabilitation program. The control group had similar inclusion criteria, with the exception for points (iii) and (iv).

The specific exclusion criteria for both the PFP and control group consisted of the following: participants should not (i) have had previous patellar dislocation or subluxation, (ii) have had previous injury or surgery to the knee, and (iii) have had recent (within the last 6 months) injury to the lower limbs (ex. Achilles tendinopathies, ankle sprain, etc.). All data collection occurred between 9^th^ January 2023 to 22^nd^ September 2023.

Ethics approval was granted by the Human Research Ethics Committee of the university (NWU-00163-22-A1), and all participants completed the informed consent forms prior to participation. An independent researcher not affiliated with the study served as an independent witness and collected the signed consent forms. All ethical procedures conformed to the requirements of ethical conduct set forth in the Declaration of Helsinki.

### Instruments

A general demographic questionnaire was used to obtain the participant’s contact details, e-mail address, age, sex, involved limb, dominant limb, previous injuries and PFP symptoms. The anterior knee pain scale (AKPS), also known as the Kujula patellofemoral scoring system, was completed electronically to capture knee-related pathologies where the total score out of 100 was captured. The AKPS was the chosen questionnaire, as it was developed specifically for evaluation of pain and disability in individuals with PFP [22], showed high test-retest reliability (intraclass correlation coefficient [ICC] = 0.95), and exhibited moderate responsiveness to clinical change, which implies that the score will likely reflect meaningful changes in a patient’s condition over time [23]. Body mass and stature were measured to calculate body mass index (BMI) [24]. Body mass was measured with an electronic scale (Seca 874, Seca, Germany) to the nearest 0.1 kg and stature with a portable stadiometer (Holtain Ltd., U.K.) to the nearest 0.01 m. A cycle ergometer (Wattbike Pro, Wattbike Ltd, Nottingham, UK) was used for a ten-minute warm-up before the testing on the ID for optimal muscle performance and reduced risk of injury. The ID (Cybex II, CSMi, Stoughton, MA, USA) was used as the gold standard for evaluating isometric muscle strength and a LC (Crane & Hanging Scale, Micro Mini CS300, Border Scales & Labels) served as the low-cost alternative to measure isometric quadriceps strength. Strength was evaluated with the knee joint at 60° of knee flexion, where peak force of the quadriceps is usually generated [25]. A digital goniometer (EasyAngle, Meloq AB, Stockholm, Sweden) was used to measure the knee angle in all instances to ensure true validity and replicability.

### Procedures

The demographic questionnaire was completed first, and only at the baseline testing. Thereafter the participants completed the AKPS questionnaire verbally, where the score was calculated out of 100 and recorded electronically. Stature and body mass were measured during barefoot standing and with minimal clothing. Lower leg length was measured from the lateral condyle of the femur to the lateral malleolus of the tibia to use as the lever length in the formula to convert force to torque for the LC measurements. Prior to testing participants completed a ten-minute warm-up on a cycle ergometer at lowest resistance and a comfortable speed (rating of perceived exertion [RPE] < 2 on the modified Borg scale). Participants completed baseline testing in two sessions separated by 24 hours, and then repeated the same testing eight weeks later. During the first session, isometric strength of the quadriceps muscles was measured first with the ID, and thereafter with the LC. During the second session, only the testing on the LC was completed for the reliability analysis. Reliability analysis was done both at baseline testing and at follow-up testing conducted after 8 weeks to ensure a stronger reliability score.

For the evaluation of isometric quadriceps strength on the ID, all variables related to the set-up were recorded to replicate the exact position for follow-up testing. The dominant limb (defined as the preferred kicking limb) in the control group and the uninjured limb in the PFP group were measured first. The participants were placed in a seated position, stabilized with upper body straps and an upper leg strap just above the knee joint. The knee was positioned at a 60° angle, using the machine angle provided by the dynamometer. The lateral femoral epicondyle was aligned with the axis of rotation of the dynamometer, and the resistance pad was positioned anterior to the distal tibia just superior to the lateral and medial malleoli. A gravity correction was performed to account for any potential additional torque induced on the attachment. The participant completed a warm-up round consisting of three repetitions, with the instruction to do one repetition at approximately 25%, 50% and 75% of perceived maximum effort before commencing of the test to assist with familiarisation of the test. A 10 second rest period was allotted between trial repetitions. A one-minute rest period was granted after the warm-up, after which the participant completed repetitions of five second maximal extension contraction with ten seconds of rest between repetitions. Verbal encouragement was given throughout the whole procedure. Pain levels were monitored by asking the participant to rate pain on a scale of 1-10 throughout the procedure (where 1=pain free, 2= very mild, 3= discomforting, 4=tolerable, 5= very distressing, 6=intense, 7=very intense, 8=utterly horrible, 9=excruciating unbearable, 10=unimaginable unspeakable) [26], and participants were permitted to stop the test if pain levels were above bearable levels (≥4). For the evaluation of isometric quadriceps torque using the LC, all positional measurements were recorded for replicability. Participants were seated on a standard chair, and the LC was attached to a strap that was attached to a fixed surface behind the chair. The strap was fastened to the participant’s ankle just above the lateral and medial malleoli where the distance between the lateral malleolus and the start of the ankle strap was measured to more accurately calculate the lever arm length in the evaluation of torque from the LC. The straps were adjusted until the knee was at 60° of flexion, measured with a digital goniometer. The participant completed the same warm-up and testing protocol as with the ID. The tests were repeated in the second session for the reliability analysis, and all procedures were repeated after 8-weeks. The conversion of raw kilogram values from the LC were converted to torque values using the following equation:

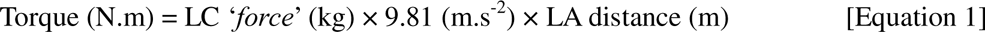

### Statistical analyses

All data were evaluated for normality using the Shapiro-Wilk test, with deviations from normality being accepted at p < 0.05. All data are presented as mean ± standard deviation (SD) unless otherwise stated. To determine the reliability of the LC, the intraclass correlation coefficient (ICC, two-way mixed effects, absolute agreement) between two measurements on separate occasions was used. The ICC values were interpreted as follows: poor: < 0.50; moderate: 0.50-0.75; good: 0.75-0.90; excellent: >0.90 [27]. Additional measures of reliability included: (i) typical error of measurement (TEM) (Equation 1), (ii) TEM% (Equation 3), and (iii) clinically meaningful difference (CMD) (Equation 4) [28]:

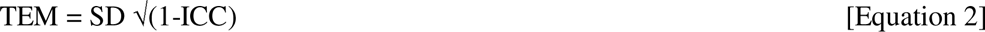

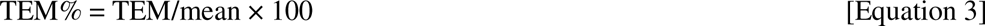

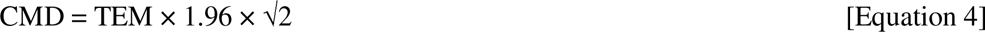

The TEM% scores were qualitatively interpreted as: excellent:<5%; good: 5-10%; poor: >10%. The CMD was used to evaluate the smallest detectable difference that would need to be overcome to conclude that a true change has occurred. A generalised linear model was used to evaluate mean differences between groups (2 levels: Con vs. PFP), and devices (2 levels: ID vs. LC) where each participant was treated as a random effect in the model. Post-hoc analyses entailed the use of paired sample t-tests with a Holm correction to adjust for multiple comparisons. Standardised mean differences were calculated as Hedge’s g, the magnitude of which was qualitatively interpreted as: trivial: <0.2; small: 0.2-0.6; moderate: 0.6-1.2; and large: > 1.2 [29].

The concurrent validity was determined by using linear regression where the Pearson correlation coefficient (r), coefficient of determination (r^2^), standard error of the estimate (SEE), and the slope of the regression lines were evaluated. The magnitude of the correlation coefficients were qualitatively interpreted as follows: negligible: 0.00-0.10; weak: 0.10-0.39; moderate: 0.40-0.69; strong: 0.70-0.89; and very strong: 0.90-1.00 [30]. Bland-Altman analyses were used to determine the systematic bias between the LC (reference measure) and the ID (criterion measure) [31]. For both the regression and Bland-Altman analyses, the point estimates were evaluated for potential outliers using Cook’s distance where potential outliers (PO) were flagged when the Cook’s distance exceeded a given threshold calculated as: 4/n (where n is the number of observations) [32]. All statistical analyses were completed using R [33].

## Results

The results pertaining to the relative (ICC_3,1_) and absolute (TEM) reliability as well as CMD of the LC are highlighted in Figure 1. Generally, the LC shows excellent relative and absolute reliability both at baseline and following the 8-week training interval. The ICC point estimates exhibit fairly narrow confidence intervals for the control group, but marginally longer intervals for the PFP group, indicating greater variability in torque measures. In all instances the CMD exceeded the TEM which implies that the LC had sufficient sensitivity to measure a true change in isometric quadricep strength (i.e. high signal-to-noise ratio).

**Figure 1:**
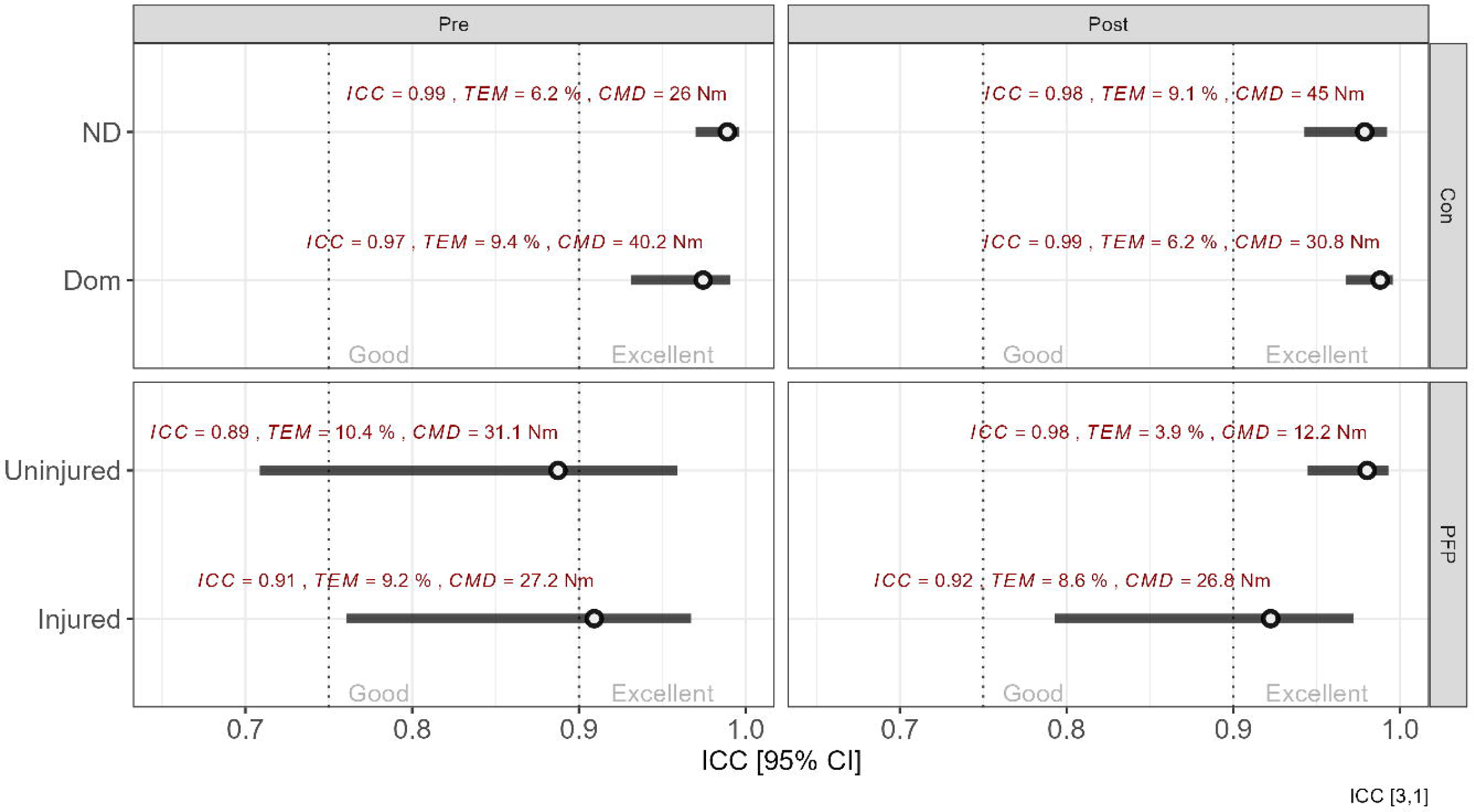
Between-group and within-group intra-class correlation coefficients. The ICC values are shown with their 95% confidence intervals. Above each point estimate are shown the precise ICC values, typical error of measurement (TEM%) and clinically meaningful difference (CMD) to provide greater context for the reliability score. Vertical dotted lines represent thresholds for excellent (ICC > 0.90) and good (ICC: 0.70-0.90). Note: ND = non-dominant; Dom = dominant; Con = control group; PFP = patellofemoral pain group.

Simple interaction effects for both between-group and within-group differences in mean torque values are shown in Figure 2. Mean differences between devices and group are shown in Figure 2A, whereas the standardised effect size with 95% CI and corresponding uncertainty density distribution for the point estimates are shown in Figure 2B.

**Figure 2:**
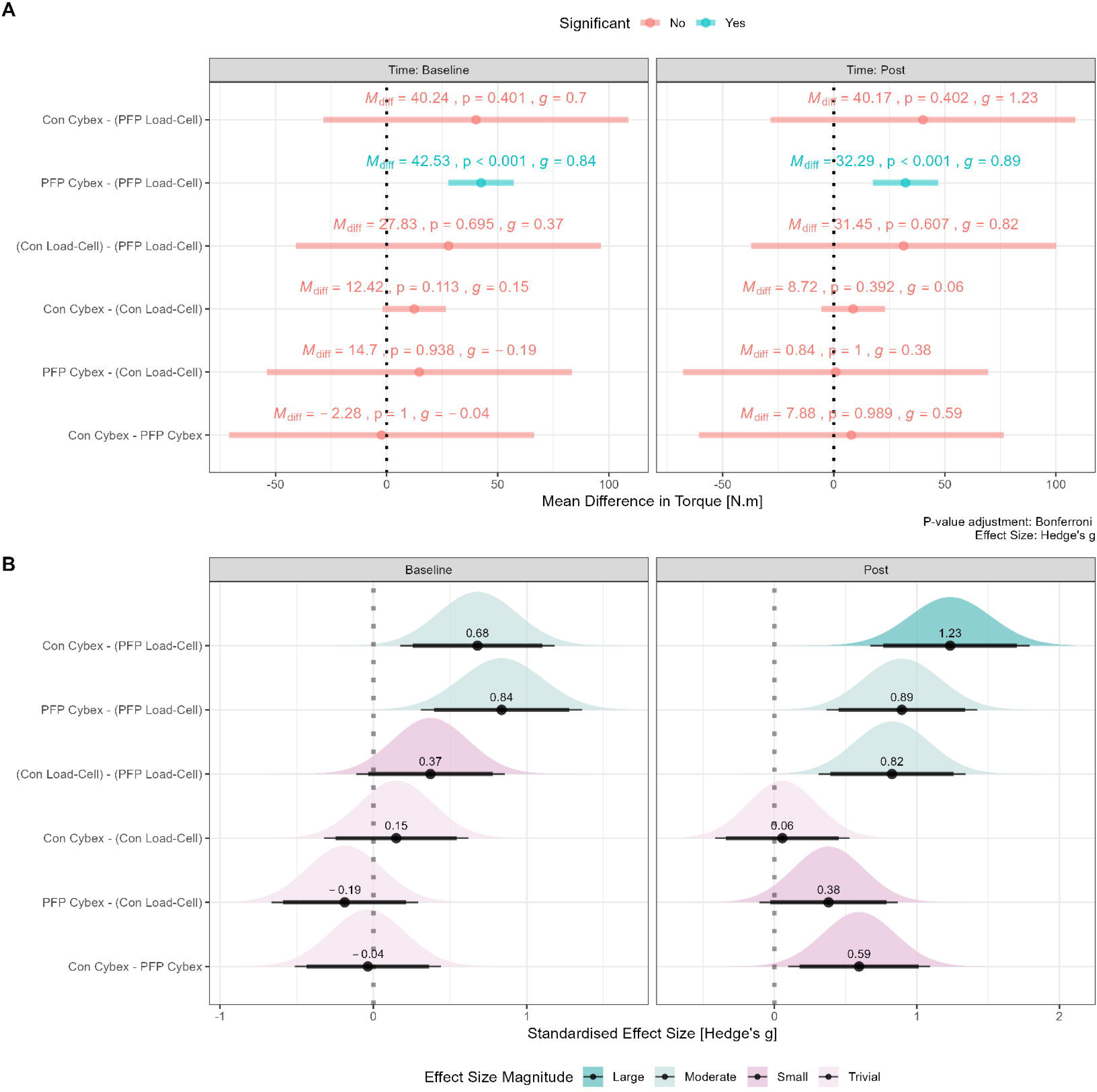
Absolute and relative within-group and between-group mean differences. Panel A: Mean differences with 95% CI; panel B: standardised mean effect size (Hedge’s g) with 90% CI (thick black line) and 95% CI (thin black line) as well as density estimates to highlight the uncertainty in the point estimate. Note: M_diff_ = mean difference; g = Hedge’s g effect size; Con = control group; PFP = patellofemoral pain group.

For the concurrent validity, strong to very strong correlations were evident between the LC and ID torque measurements (r = 0.88-0.90, SEE = 0.05-0.07 Nm) (see Figure 3). Based on the slope analysis however, it is important to note that the torque values from the LC are typically lower than that obtained from the ID (slope = 0.65-0.77).

**Figure 3:**
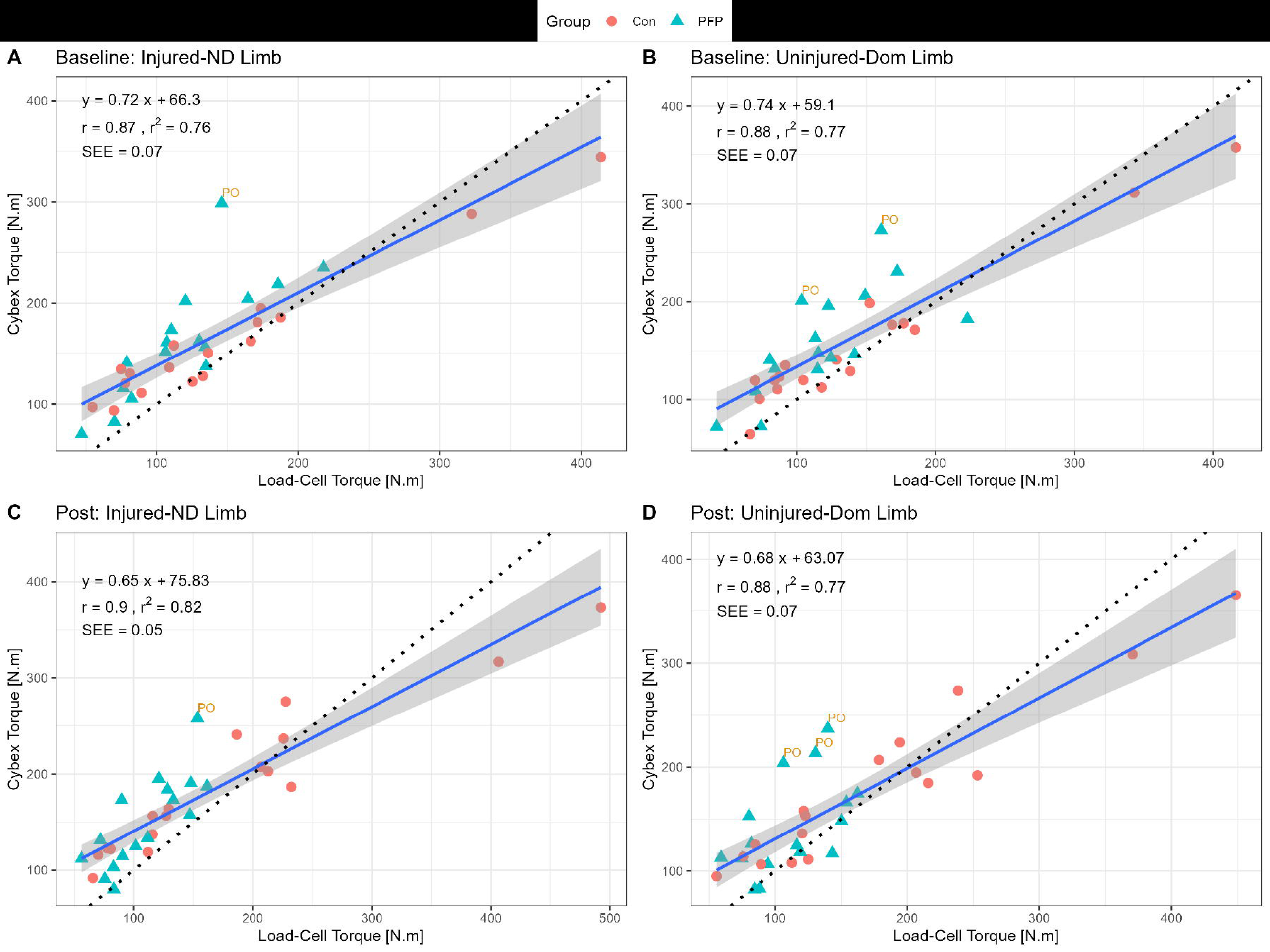
Linear regression between load-cell and Cybex for isometric knee extension torque. PO = potential outlier; SEE = standard error of the estimate; ND = non-dominant; Dom = dominant

The results from the Bland-Altman analysis confirm the measurement bias between the LC and the ID whereby an average systematic bias of 16.3-28.8 Nm is evident in favour of the ID (see Figure 4).

**Figure 4:**
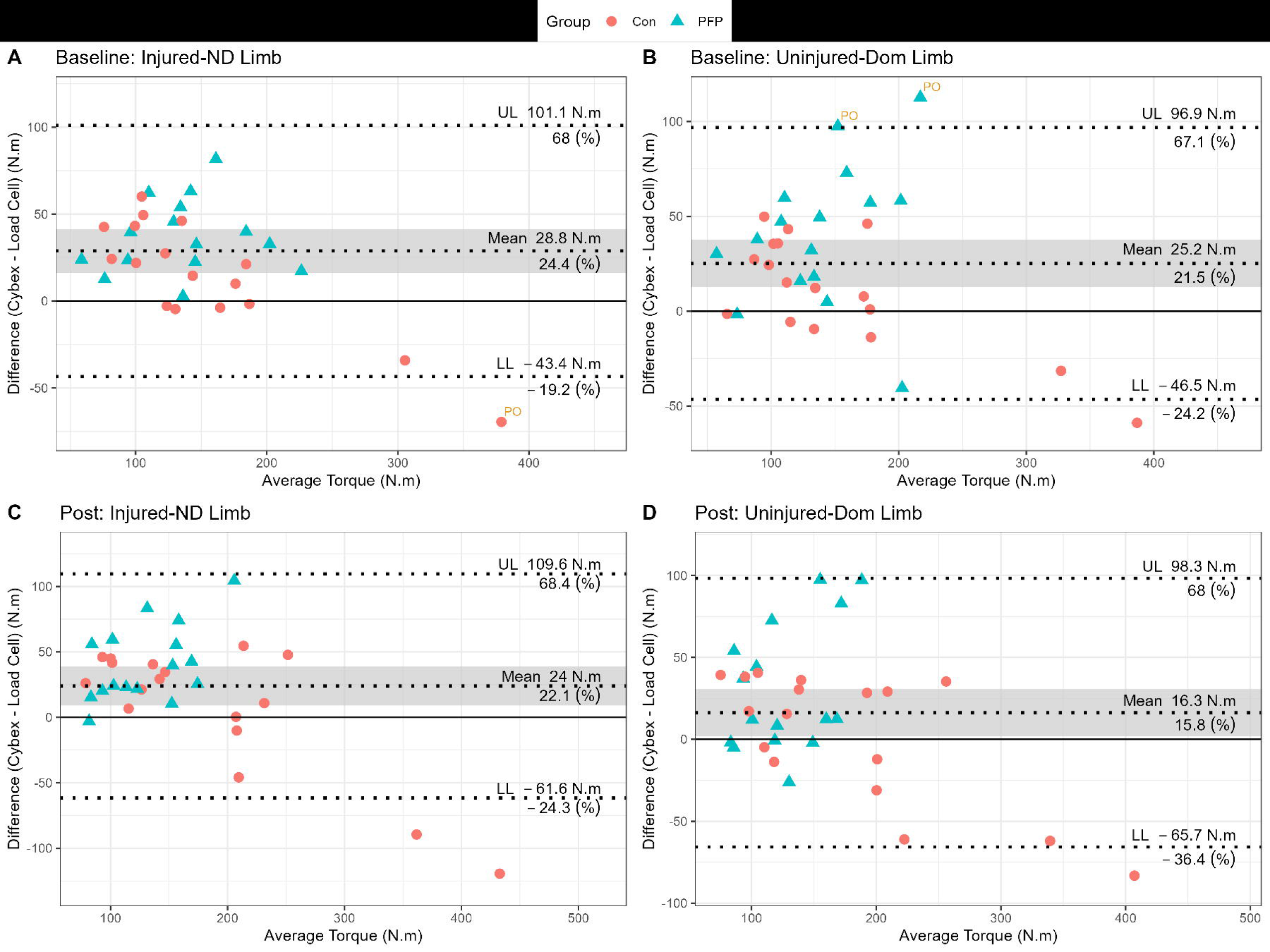
Bland-Altman plots for the bias (95% CI) between instruments and the limits of agreement. Note: UL = upper limit; LL = lower limit; ND = non-dominant; Dom = dominant; PO = potential outlier

Within-group differences in peak torque for each group and each device are shown in Figure 5. Although no statistically significant differences are evident, it is important to emphasise the inter-individual variability highlighted by the colour-gradients which indicate the magnitude of participant-specific torque improvements in quadriceps strength between time points.

**Figure 5:**
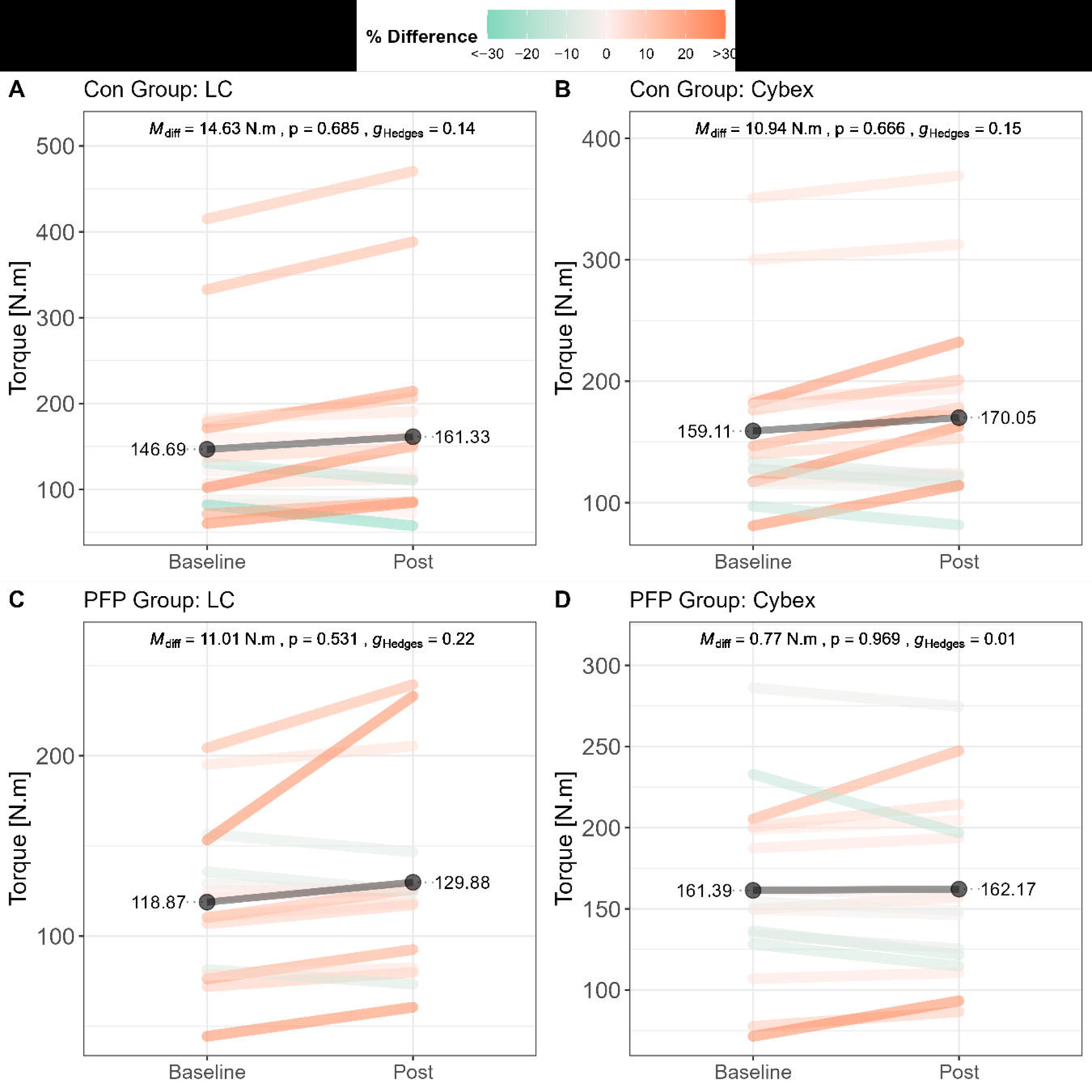
Within-group differences in peak torque. Pre-post differences are shown for the control group as evaluated by the LC (panel A) and ID (panel B). Pre-post differences are shown for the PFP group as evaluated by the LC (panel C) and ID (panel D). Line segments are coloured based on the percentage difference (Post-Pre/Pre×100) where improvements are shifted towards orange, and decrements are shifted towards green. The magnitude of the difference is captured by the intensity of the colour gradient (larger differences are darker, smaller differences are lighter). Black lines indicate the group mean for a given time point. Note: Mdiff = mean difference; g_Hedges_ = Hedge’s g; Con = control group; PFP = patellofemoral pain group; LC = Load Cell

## Discussion

The present study yielded several novel findings. Firstly, the reliability and validity of a low-cost, commercially available LC were evaluated in comparison to a gold standard ID. The utility of such a device would have meaningful implications in rehabilitation services, especially in under-resourced practices. Secondly, we showed that the LC exhibited exceptional sensitivity for detection of clinically meaningful changes in isometric knee torque. Finally, we evaluated differences in isometric knee torque between a control and PFP group following an 8-week period showcasing substantial within-subject variability and constrained between-group differences.

### Reliability

The LC showed excellent relative and absolute reliability in both the control and PFP groups (ICC>0.90 and TEM <10%) in all instances apart from the uninjured limb of the PFP group. The large ICC values obtained in the present study compare favourably with those from a separate study where belt-stabilisation was used with an HHD (ICC = 0.62 to 0.96) [34]. Similarly, the TEM% scores of the present study were substantially better those reported by Martins *et al.* [34] who recorded TEM% of 12% for a belt-stabilised HHD and Chamorro *et al.* [18] who yielded values ranging between 4-15% for measurements without stabilisation, both for knee extension strength. The ICC point estimates showed narrow confidence intervals in which the upper and lower limits fell within the margin defined as *excellent* in most instances. However, longer confidence intervals are observed in the PFP group than in the control group at the initial set of testing, indicating that the PFP group had greater variability in torque measures in the injured as well as the uninjured limbs compared to the control group. Torque variability for knee extensors is present in other knee injuries as well [35], and in this case the variability may be accounted for in the PFP group on the basis that PFP patients tend to have impaired quadriceps function often ascribed to impaired vastus medialis oblique firing [36,37] and reduced eccentric control [38]. However, the variability improved notably in the uninjured limb following the intervention period, possibly due to enhanced motor learning which tends to occur in injured populations [39].

It should be noted that the reliability of the LC, as with any device, is dependent on the set-up and therefor clinicians should be vigilant in following the set-up instructions set out in the procedures section and keep the set-up consistent when doing testing and re-testing in clinical settings, to ensure reliable results. Belt-stabilisation seems to be improving reliability in LC-based devices and are recommended to use by practitioners [25,34].

### Clinically meaningful difference

The CMD within the present study ranged between 12-45 Nm, implying that the differences in torque production by the quadriceps musculature between limbs or over the course of rehabilitation can be detected with the LC. Previous research has shown that an HHD exhibited comparable, although marginally lower CMD values of 17-27 Nm [40], with the ID yielding the lowest and most consistent CMD of approximately 27 Nm [41].

### Group differences in torque production

An intriguing finding of the present study relates to the simple interaction effects regarding within-group (e.g., ID vs. LC) and between-group (e.g., Control vs. PFP) differences for quadriceps torque (see Figure 2A, 2B). The mean differences in torque production between devices and groups are classified as trivial-to-moderate, and typically range −2.28 Nm to 27.83 Nm (p = 0.113-0.999). In most instances the point estimates of the mean difference were not significantly different from zero largely due to the longer confidence intervals. Such a result generally implies, at least in principle, similarities in torque production between groups and between devices. The standardised effect sizes (together with the 95% CI) show that, despite a lack of statistical significance, the magnitude of the mean differences are likely to be meaningful, especially for the PFP group which consistently yielded higher torque values on the ID compared to the LC (M_diff_ = 43.53 Nm, p < 0.001). The interaction effects should be of interest to practitioners on the basis that the interchangeability of device measurements must be considered when interpreting results and making clinical inferences [41]. Given that, at least in some instances, practitioners might receive isokinetic results for a specific PFP patient or a referring clinician, these results should be verified when using the LC in the clinical setting so that more precise interpretations can be made when evaluating temporal changes in quadriceps strength. Moreover, the same device should be used by the same clinician to ensure adequate consistency in readings and insights of whether true changes have accrued.

### Validity

The results of the preset study suggest that the concurrent validity of the LC, when compared to the ID, appears to be strong to very strong, although a few discrepancies are noteworthy. The torque values from the LC are typically lower than those from the ID, with the exception of torque values exceeding ∼300 Nm (see Figure 3 and Figure 4). It is therefore reasonable to state that the LC might be of better use in injured and non-athletic populations that will most likely produce lower torque values compared to healthy, uninjured, or very athletic participants that might produce substantially higher torque values. The validity of the LC was greater than that of an HHD which yielded poorer scores even when stabilised with a belt, with correlations ranging between r = 0.3-0.8 [18,42]. The LC is not a perfectly valid tool given the bias in torque readings and moderate-to-large limits of agreement (LoA) and is therefore unlikely to replace the ID for absolute values. More specifically, there is evidence of a systematic bias in favour of the ID (see Figure 4), confirming that the LC typically measures lower torques than the ID on average. These results correspond with Martins et al. [34], who showed that knee extension tested with a belt-stabilised HHD, exhibited similar mean difference in torque production, with bias towards the ID. The HHD also shows, on average, wide LoA values (33.59%, CI_95%_ [23.91%, 43.26%]) for knee extension in other literature [18]. The wide LoA therefore support the notion that the LC is not valid enough to replace values obtained from the ID but given the excellent reliability would still be an exceptional tool for the evaluation of isometric quadriceps strength. It is also important to highlight that isokinetic norms should not be used to make clinical interpretations when comparing these to values derived from the LC. It would be important to develop independently generated normative data for LC-derived torque values to facilitate decision-making across different joints and population groups.

### Temporal changes in torque

Finally, although the mean peak torque values did not change significantly from the baseline to follow-up testing (M_diff_ = 0.77-14.63 Nm, p = 0.531-0.969), it should be noted that there were substantial individual improvements both within- and between groups (see Figure 5). The variability in individual responses underscores the potential inadequacy of the intervention programmes followed by the groups which were beyond the control of the current study. Whether more focused and targeted interventions or prolonged rehabilitation timelines (> 8 weeks) would elicit more favourable outcomes would require further research, especially in those with PFP. It should however be noted that there were consistencies in the measured values between devices across time, implying that the LC indeed has exceptional utility as a measurement tool for evaluating isometric quadriceps strength.

### Limitations

Given the strengths of the present study, it is also important to underscore some limitations. Firstly, participants could follow any rehabilitation program at any practitioner of their choice, and therefore could not control the details associated with targeted quadriceps strengthening as this was beyond the scope of the present study. Secondly, the ratios of males versus females in the groups differed, although consistency within groups were more important for this study than consistency between groups. Thirdly, participants were not divided into different groups according to the magnitude of quadriceps strength deficits. This latter point could provide more nuanced information on strength improvements over time in future studies.

## Conclusions

The objectives of this study were to determine the validity and reliability of a low-cost LC for use in a clinical setting for measuring knee extension strength in those with PFP. The LC exhibited excellent reliability, and was deemed sensitive enough to detect clinically meaningful differences over time in both healthy individuals as well as those with PFP. The reliability of the LC is dependent on the set-up of the individual, and therefore practitioners should take care to complete the set-up as described and ensure consistency across every testing session. The LC lacks validity and is therefore unlikely to be an adequate surrogate for isokinetic dynamometry. However, given the trade-off in (i) costing associated with the ID and the LC, (ii) the importance of evaluating and tracking changes in knee extension strength in those with PFP, and (iii) the excellent reliability of the LC, the utility of the LC as a viable assessment tool is advocated, especially in resource-restricted settings.

## Data Availability

https://doi.org/10.7910/DVN/N1SSPT

